# Persistence of SARS-CoV-2 omicron variant in children and utility of rapid antigen testing as an indicator of culturable virus

**DOI:** 10.1101/2022.06.21.22276668

**Authors:** Zoe M. Lohse, Jerne J. Shapiro, John A. Lednicky, Melanie N. Cash, Inyoung Jun, Carla N. Mavian, Massimiliano S. Tagliamonte, Cyrus Saleem, Yang Yang, Eric J. Nelson, Marco Salemi, Kathleen A. Ryan, J. Glenn Morris

**Author notes:** Corresponding author:* J. Glenn Morris, Jr., Emerging Pathogens Institute, University of Florida. Alternative corresponding author:* John A. Lednicky, Emerging Pathogens Institute, University of Florida.

## Abstract

We screened 65 longitudinally-collected nasal swab samples from 31 children aged 0-16 years who were positive for SARS-CoV-2 omicron BA.1. By day 7 after onset of symptoms 48% of children remained positive by rapid antigen test. In a sample subset we found 100% correlation between antigen test results and virus culture.

---

The Coronavirus disease 2019 (COVID-19) pandemic began with the emergence of the novel coronavirus SARS-CoV-2 in late 2019. Since the first reported case of COVID-19 in China, the intensity of the pandemic and our understanding of disease transmission has ebbed and flowed with the introduction of competing variants worldwide [1].

On July 20, 2020, the U.S. Centers for Disease Control and Prevention (CDC) recommended that persons infected with SARS-CoV-2 isolate for a 10-day period, without requirements for an associated negative test for the virus [2]. This followed from studies showing that virtually all non-immunosuppressed patients with mild to moderate disease became culture-negative by day 10 of infection, with virus culture providing what was regarded as the optimal laboratory marker for infectivity [3-5]. The Florida Department of Health (FDOH), in an Emergency Rule issued September 21, 2021 [6] indicated that children infected with SARS-CoV-2 could return to school sooner than 10 days if they had a negative COVID test and were asymptomatic. On January 14, 2022, CDC changed their recommendation to say that children and adults infected with SARS-CoV-2 should isolate for 5 days, and, if afebrile, could then return to work/school but should wear a high-quality mask for an additional 5 days [7,8]. The FDOH position was changed on February 24, 2022, to say that children only needed to isolate for 5 days before returning to school, without further testing, and that masking was not necessary [9]. In this somewhat confusing setting, as the Omicron variant was spreading rapidly in Florida, we sought to characterize SARS-CoV-2 Omicron persistence and infectivity among children with mild illness in an outpatient setting.

Children with a positive rapid antigen test or a PCR-based test for SARS-CoV-2 were referred from the UFHealth pediatric clinic or by school nurses at a near-by local public school [10]. We focused on enrolling children who were five or more days from onset of symptoms, with samples collected between 12/20/2021 and 2/21/2022. In two instances in which multiple children from a family were enrolled, we also collected samples from parents of the participating children. The study was approved by the University of Florida IRB (protocol IRB202000488), with signed informed consent obtained from participants. Parents consented for children under the age of 18, with assent for children over seven years of age.

We obtained 65 nasal swab samples from 31 children aged 0-16 years, and eight samples from three parents. Two anterior nasal swab samples were obtained from each participant: one was immediately tested with the BinaxNOW rapid antigen test (Abbott Laboratories, Abbott Park, Ill), and the second was frozen at -80°C for subsequent culture, reserve transcriptase quantitative polymerase chain reaction (RT-qPCR), viral load determination and sequencing. Children who had a positive result on the initial follow-up test done as part of this study were asked to return on day 7, 10, and thereafter, with testing continuing until a negative test result was obtained. If a negative rapid antigen test result was obtained before day 10 of isolation, children were given a note which permitted them to return to school, as specified in the September 2021 FDOH Emergency Rule.

BinaxNOW testing was done following the manufacturer’s instructions, and participants were provided with results as soon as testing was completed. As described in Supplementary Methods, RT-qPCR [11] was performed and a standard curve generated using N1 quantitative standards 10-fold diluted to determine viral copies. cDNA synthesis and library preparation were performed using the COVIDSeq Test kit and Mosquito HV Genomics Liquid Handler. Sequences were aligned with those from other local cases to determine relatedness with community spread using viralMSA and the MN908947 reference sequence [12]. A maximum likelihood phylogenetic tree was reconstructed using IQ-TREE with the best fitting nucleotide substitution model according to the Bayesian Information Criterion and 1,000 bootstrap replicates [13].

We used a variety of cell lines for virus culture, including LLC-MK2 and Vero E6 cells and A549 cells expressing ACE-2, HEK 293T cells expressing human ACE-2, and VeroE6 with high endogenous ACE-2 [14-16]; see supplementary methods for details. Cells were observed daily for one month before being scored negative for virus isolation. When virus-induced cytopathic effects (CPE) were evident, the presence of SARS-CoV-2 was determined by RT-PCR [17,18]. Isolation of SARS-CoV-2 at or after six days post-inoculation of cells was most effective in VeroE6-ACE-2 and HEKT293-ACE-2 cells (10^4^ to 10^7^ genome equivalents/µL of purified vRNA), with marginally lower virus yields in LLC-MK2 and VEROE6 cells, and generally low yields in A549-ACE-2 cells (10^1^ to 10^2^ genome equivalents/µL of purified vRNA).

Median age of the 31 children enrolled was seven years (range 0-16), including 10 children under the age of five; two-thirds were boys. Results of the rapid antigen test were negative for 10 children at the time they enrolled in our study, which was day 4 or later after onset of symptoms and/or their initial positive test for SARS-CoV-2; these 10 children were not tested further. We obtained at least one positive rapid antigen test for the remaining 21 children, with a subsequent negative test for 14 children, collected a median of 2 days after their last positive test. Including the 10 children who had an initial negative test and assuming that children would be positive on all days before their last positive test, 67% would have been positive for SARS-CoV-2 on day 5, with 48% still positive by rapid antigen on day 7 (Figure 1). We fitted a parametric survival model to estimate the percent of remaining positive over time, accounting for interval-censoring and right-censoring of the exact transition times from positive to negative (see supplementary methods). The mean and median durations of remaining positive were estimated to be 7.74 (95% CI: 6.54, 9.17) days and 7.51 (95% CI: 6.24, 9.04) days, respectively, and the inter-quartile range is (5.23, 9.99)(Supplementary Figure S1). The model predicted that the probability of remaining positive dropped to 10% by day 12, close to our data-based estimate of the percent positive.

**Figure 1:**
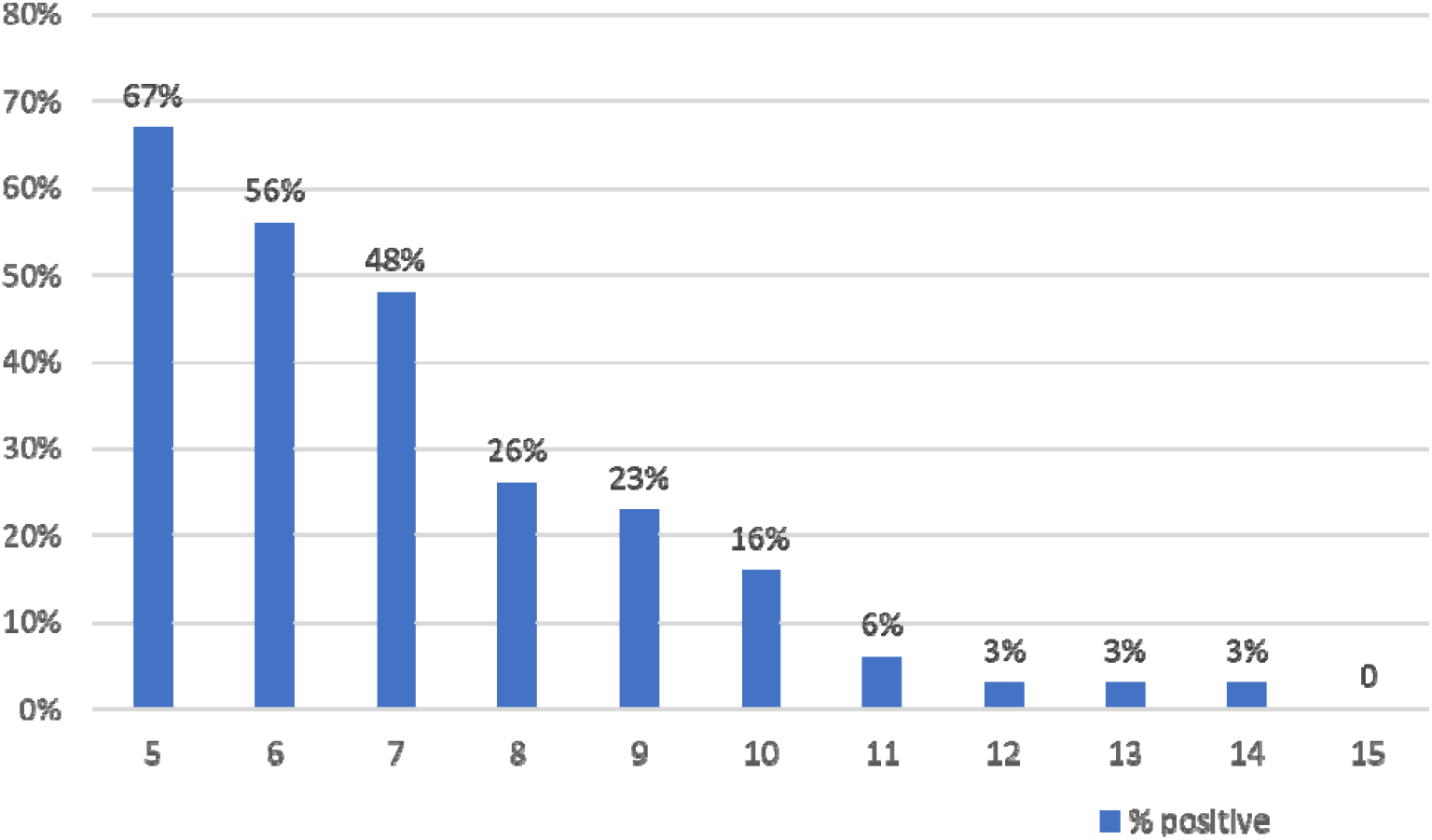
Percent of children with positive rapid antigen test, by day of infection

Virus cultures were performed for the first 15 samples collected from children and the eight samples from parents. SARS-CoV-2 grew in cultures of 16 of these 23 samples (11 from children, five from adults), all of which were also positive by rapid antigen testing. No growth was detected in seven cultures (four samples from children, three from adults), all of which were negative by rapid antigen testing. The correlation between culture and rapid antigen testing results was significant for both children and adults (p<0.0001 for children, p=0.02 for adults, Fisher’s Exact).

A total of 40 positive rapid antigen test results were obtained for children enrolled in the study; all were positive by RT-qPCR. Among 25 samples with a negative rapid antigen test result, 9 were RT-qPCR positive, with a median viral load of log_10_ 3.45 copies/mL (range log_10_ 3.08-4.23 copies/mL). Four of these nine samples had been cultured; all were culture-negative. For children for whom serial samples were available, all showed a consistent pattern of decreasing viral load across time. We did not see an age-related difference in viral load after correcting for day of illness, nor was viral load or duration of infection correlated with vaccination status (42% of children were vaccinated) [19]. Representative results from one child for whom eight serial samples were available are shown in Table 1: this was an otherwise healthy child under the age of five years who was febrile (38.3°C) with mild upper respiratory symptoms for one day when first diagnosed, with no further symptoms.

**Table 1:** Results of SARS-CoV-2 rapid antigen testing, cell culture, cycle times (CT value), viral load (log10 copies/mL) and symptoms, by days of infection in otherwise healthy child under the age of five years.

| Day of Infection | Rapid antigen testing | Cell culture | CT value | Viral load, (Log<sub>10</sub> copies/mL) | symptoms |
| --- | --- | --- | --- | --- | --- |
| 6 | pos | pos | 22.12 | 8.5 | none |
| 8 | pos | pos | 24.04 | 7.6 | none |
| 10 | pos | pos | 31.15 | 5.8 | none |
| 11 | pos | pos | 30.48 | 5.5 | none |
| 12 | pos | pos | 33.95 | 4.7 | none |
| 13 | pos | pos | 35.20 | 4.6 | none |
| 14 | pos | pos | 33.61 | 4.2 | none |
| 16 | neg | neg | 38.10 | 3.3 | none |

All SARS-coV-2 positive samples were sequenced, and all were found to be omicron BA.1; sequence data have been submitted to GISAID (see supplementary material). Phylogenetic analysis revealed minimal variability across the sample set, consistent with spread of a single clade within the community. While approximately one-third of children in the sample attended a single public school, we did not see any evidence of increased clustering among children from that school. We did identify tighter clustering within families (including the families for which we had sequence data for both children and parents) as might be expected if transmission was occurring primarily within families rather than within community settings.

## Comment

While the numbers of participants and samples analyzed were low, we found an exact correlation between results from the BinaxNOW rapid antigen test and results of cell culture, generally accepted as the best marker for virus infectivity [3-5,19]. We did have samples that were negative by rapid test and positive by rRT-PCR; however, viral loads in these instances were low, and it is unclear that children from whom these samples were obtained would have been infectious, particularly in light of the negative cultures obtained. We emphasize that these findings are specific for SARS-CoV-2 omicron BA.1, and it will require epidemiologic studies to fully assess infectivity; results may well differ with other variants. We note that all samples for this study were obtained on day 4 or thereafter, so we cannot comment on possible delays in obtaining an initial positive result with rapid antigen testing.

In this study close to half of infected children remained positive for SARS-CoV-2 (and likely were infectious) for at least two days after they would have returned to school under the January 14, 2022 CDC guidelines. While transmission would be reduced by wearing a mask, as recommended by CDC, wearing of masks in school settings was highly controversial in Florida, and FDOH specifically recommended against the use of masks in their February 24, 2022 statement [9]. There is clear value, from a social standpoint, in minimizing the isolation period of children infected with SARS-CoV-2. At the same time, from a public health standpoint, there is a need to minimize the risk that infected children will continue to transmit the virus after their return to school. Consideration should be given to lengthening the current recommendation for five days of isolation before return to school, potentially in combination with requirements for a negative rapid test result.

## Data Availability

All data produced in the present study are available upon reasonable request to the authors, contingent upon compliance with requirements of the University of Florida IRB

---

**Supplementary Figure S1.**
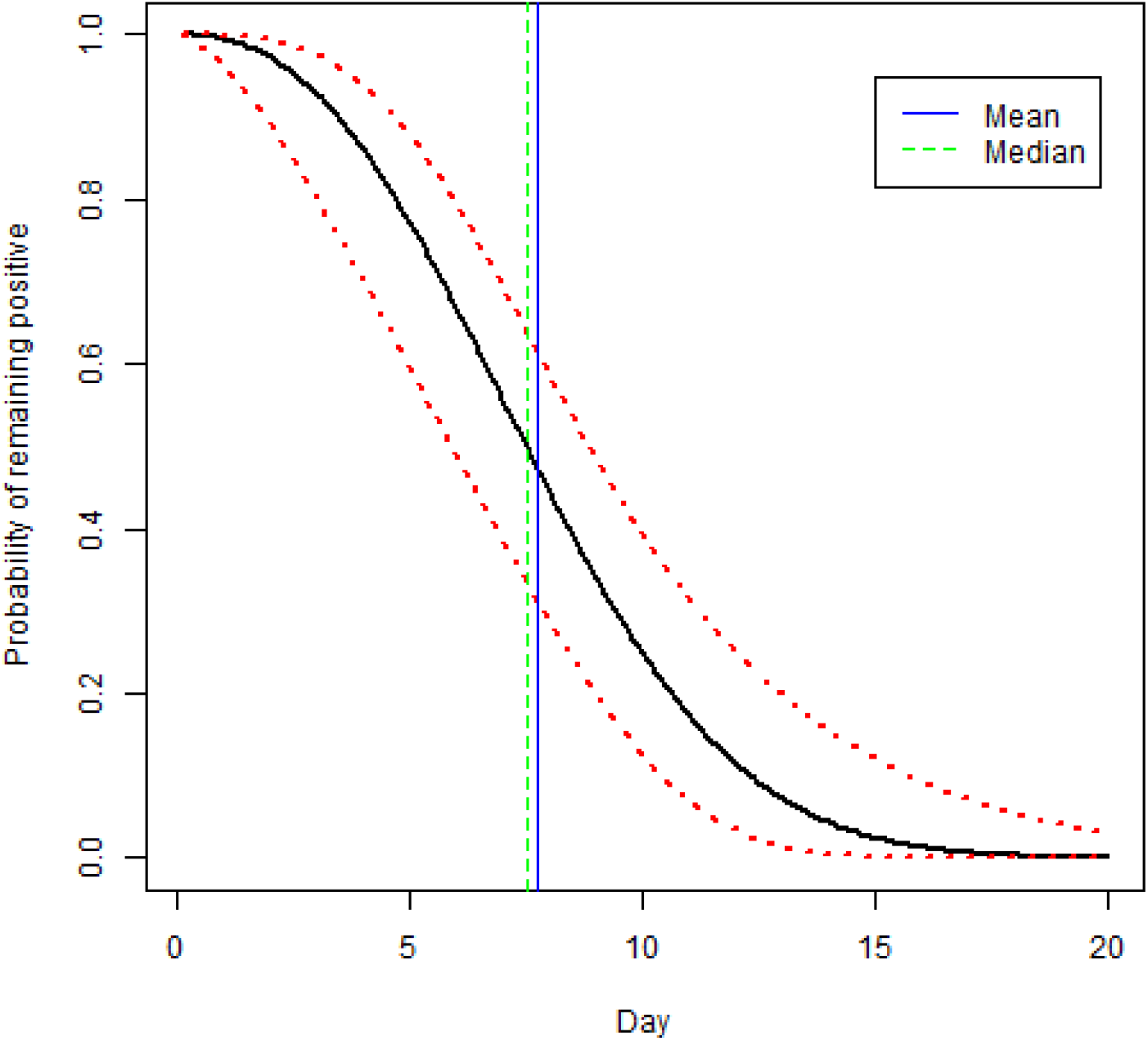
Model-predicted probability of remaining test-positive since symptom onset. This probability curve (black solid) is simply the survival function for the fitted Weibull model, and the 95% confidence bands (red dashed) are derived using the delta method. Mean (blue solid) and median (green dashed) times from symptom onset to turning negative are shown as vertical lines.

## REFERENCES

1. CDC. About Variants. https://www.cdc.gov/coronavirus/2019-ncov/variants/about-variants.html?s_cid=11723:covid%2019%20variants%20of%20concern:sem.ga:p:RG:GM:gen:PTN:FY22. Accessed 6/2/2022; page last updated 4/26/2022.

2. CDC. Updates from previous ending home isolation webpage content. https://www.cdc.gov/coronavirus/2019-ncov/hcp/duration-isolation.html#anchor_1631644826603. Accessed 6/13/2022.

3. Walsh KA, Spillane S, Comber L, et al. The duration of infectiousness of individuals infected with SARS-CoV-2. J Infect 2020;81:847–856. https://doi.org/10.1016/j.jinf.2020.10.009.

4. Cui C, Kweon O-J, Jung S-Y, et al. Duration of culturable SARS-CoV-2 in hospitalized patients with COVID-19. N Eng J Med 384;7 DOI:10.1056/NEJMc2027040

5. Owusu D, Pomeroy MA, Lewis NM, et al. Persistent SARS-CoV-2 RNA shedding without evidence of infectiousness: A cohort study of individuals with COVID-19. J Infect Dis 2021;224:1362–71. DOI:10.1093/infdis/jiab107.

6. CDC. Overview of COVID-19 Isolation for K-12 Schools. https://www.cdc.gov/coronavirus/2019-ncov/community/schools-childcare/k-12-contact-tracing/about-isolation.html. Accessed 6/2/1022; page last updated 1/6/2022.

7. CDC. Ending Isolation and Precautions for People with COVID-19: Interim Guidance. www.cdc.gov/coronavirus/2019-ncov/hcp/duration-isolation.html. Accessed 6/2/2022; page last updated 1/14/2022.

8. Florida Department of Health. Notice of Emergency Rule. Rule No:64DER21-12, Protocols for controlling COVID-19 in school settings, August 6, 2021.

9. Florida Department of Health. COVID-19 Guidance Recommendations, February 24, 2022.

10. Nelson EJ, McKune SL, Ryan KA, et al. Antigen versus RT-PCR tests for screening quarantined students during a SARS-CoV-2 delta variant surge. JAMA Pediatr. Published online March 7, 2022. doi:10.1001/jamapediatrics.2022.0080

11. CDC - National Center for Immunization and Respiratory Diseases (NCIRD), Division of Viral Diseases - https://www.cdc.gov/coronavirus/2019-ncov/lab/rt-pcr-panel-primer-probes.html/. June 6, 2020.

12. Moshiri N. ViralMSA: Massively scalable reference-guided multiple sequence alignment of viral genomes. Bioinformatics 2020. doi:10.1093/bioinformatics/btaa743.

13. Nguyen LT, Schmidt HA, von Haeseler A, Minh BQ. IQ-TREE: a fast and effective stochastic algorithm for estimating maximum-likelihood phylogenies. Mol Biol Evol 2015;32:268–274, doi:10.1093/molbev/msu300.

14. Zhao H, Lu L, Peng Z, et al. SARS-CoV-2 Omicron variant shows less efficient replication and fusion activity when compared with Delta variant in TMPRSS2-expressed cells, Emerging Microbes & Infections 2022;11(1):277–283, DOI:10.1080/22221751.2021.2023329

15. Yadav PD, Gupta N, Potdar V, et al. Isolation and Genomic Characterization of SARS-CoV-2 Omicron Variant Obtained from Human Clinical Specimens. Viruses 2022;14(3):461. https://doi.org/10.3390/v14030461

16. Suzuki R, Yamasoba D, Kimura I et al. Attenuated fusogenicity and pathogenicity of SARS-CoV-2 Omicron variant. Nature (2022). https://doi.org/10.1038/s41586-022-04462-1

17. Lednicky JA, Lauzard M, Fan ZH, et al. Viable SARS-CoV-2 in the air of a hospital room with COVID-19 patients. Int J Infect Dis. 2020 Nov;100:476–482. doi: 10.1016/j.ijid.2020.09.025. Epub 2020 Sep 16. PMID: 32949774; PMCID: PMC7493737.

18. Lednicky JA, Shankar SN, Elbadry MA, et al. Collection of SARS-CoV-2 virus from the air of a clinic within a university student health care center and analyses of the viral genome. Aerosol Air Qual Res 20 (2020), pp. 1167–1171.

19. Boucau J, Marino C, Regan J, et al. Duration of viable virus shedding in SARS-CoV-2 omicron variant infection (p. 2022.03.01.22271582). medRxiv. https://doi.org/10.1101/2022.03.01.22271582

