## Supplementary Methods for "Persistence of SARS-CoV-2 omicron variant in children and utility of rapid antigen testing as an indicator of culturable virus"

***Sample Collection/Rapid Antigen Testing:*** Abbott BinaxNOW testing was done by trained public health students following the manufacturer’s instructions. Anterior nasal swab samples were obtained following the procedure outlined in the test kit and using the swab from the test kit. Participants were provided with results of the rapid antigen test as soon as testing was completed. Following CDC definitions, day 0 of infection was defined as the day of onset of symptoms or the day a positive test for SARS-CoV-2 (PCR or rapid test) was obtained, whichever came first.

Using the same pattern of swabbing as done for the BinaxNOW testing, a second swab (Coventry sterile nylon sampling swabs, Chemtronics, Kennesaw, Ga) was collected from each participant and placed in 1 ml of virus transport media. A customized virus transport media (VTM) was used for this study: it was formulated with cryoprotectants in the form of additional protein and sucrose to preserve infectivity of the virus. It is comprised of 1 × brain heart infusion broth (Difco, US) supplemented with 0.4 mg/mL neomycin [sulfate](https://www.sciencedirect.com/topics/earth-and-planetary-sciences/sulphate) (Cell culture grade, CAS Number: 1405-10-3, Sigma Aldrich, US), 2.5 μg/mL gentamycin (Cell culture, CAS no. 1405-41-0, Sigma Aldrich, US), 0.2 M sucrose (Cell culture grade, CAS no. 57-50-1, Sigma Aldrich, US) and 4 g/L of bovine serum [albumin](https://www.sciencedirect.com/topics/earth-and-planetary-sciences/albumin) (BSA) fraction V (Lyophilized, catalog no. 15260037, Sigma Aldrich, US)[1]. After collection, samples were stored at -80°C in a continuously monitored freezer.

**PCR/Viral Load/Sequencing:** For processing, nasal swab samples were thawed for 10 minutes at room temperature, the swab in each sample mixed thoroughly with the VTM using BSL2+ work practices, and the resultant sample was divided into aliquots of ca. 200 ul. Viral RNA was extracted from the sample by using the QIAamp 96 Viral RNA Kit according to the manufacturer’s instructions with the QIAcube HT (Qiagen, Germantown, MD) using the following settings with a filter plate: the lysed sample was premixed 8 times before subjecting to vacuum for 5 minutes at 25kP and vacuum for 3 min at 70kPa. Following 3 washes using the same vacuum conditions above, the samples were eluted in 75 µl AVE buffer followed by a final vacuum for 6 minutes at 60 kPa.

Levels of SARS-CoV-2 RNA were determined using the 2019-nCoV_N1 assay (primer and probe set) with 2019-nCoV_N_positive control (IDT, Coralville, Iowa) per CDC guidelines [2]. Viral RNA was subjected to first-strand synthesis using ProtoScript II Reverse Transcriptase according to the manufacturer’s instructions (New England Biolabs, Ipswich, MA). Quantitative PCR was performed using TaqMan Fast Advanced Master Mix (ThermoFisher Scientific, Waltham, MA) according to the manufacturer’s instructions. A standard curve was generated using N1 quantitative standards 10-fold diluted to determine viral copies. The assay was run in triplicate including a non-template control [3]. Viral copies were estimated from Ct values and demonstrated here as the logarithm base 10 using the following formula: Copy number = 10^(Ct - Intercept)/(Slope).

cDNA synthesis and library preparation were performed using the COVIDSeq Test kit (Illumina, San Diego, CA) and Mosquito HV Genomics Liquid Handler (SPT Labtech Inc., Boston MA). Libraries were sequenced using the NovaSeq 6000 Sequencing System SP Reagent Kit and the NovaSeq Xp 2-Lane Kit. Illumina’s DRAGEN pipeline was used to derive sample consensus sequences, which were filtered based on a minimum of 70% coverage of the genome. Sequences were aligned together with the sequences from other local cases available in our in-house Florida database to determine relatedness with community spread using viralMSA and the MN908947 reference sequence [4]. Mutations potentially associated with contamination, recurrent sequencing errors, or hypermutability were masked using a vcf filter (<https://virological.org/t/masking-strategies-for-sars-cov-2-alignments/480>). Lineages for all sequences were determined using the PangoLEARN model (Pangolin v 2.3.6)[5]. A maximum likelihood phylogenetic tree was reconstructed using IQ-TREE with the best fitting nucleotide substitution model according to the Bayesian Information Criterion (BIC) and 1,000 bootstrap replicates [6].

All sequence data have been submitted to GISAID. Accession numbers include 13288898-902, 13288909, 13288968-975, 13288977-979, 13288981-989, 13288992-994, 13291615, and 13291623.

***Cell culture/Virus isolation:*** Susceptible and permissive cells that express human angiotensin-converting enzyme 2 (ACE-2) and or transmembrane serine protease 2 (TRMPSS-2) have typically been used for the isolation of SARS-CoV-2 omicron variants, exemplified by the reports of Zhao *et al*., Yadav *et al*., and Suzuki *et al*. [7-9]. Isolation was therefore attempted using cell lines obtained from the American Type Culture Collection (ATCC): this included LLC-MK2 (Rhesus monkey kidney cells, catalog no. ATCC CCL-7) and Vero E6 cells (African green monkey kidney cells, catalog no. ATCC CRL-1586) [10], and also, A549 cells expressing ACE-2 (ATCC NR-53726), HEK 293T cells expressing human ACE-2 (ATCC NR-52511), and VeroE6 high endogenous ACE-2 (ATCC NR-53726). The cells were propagated in cell culture medium comprised of aDMEM (advanced Dulbecco’s modified essential medium, Invitrogen, Carlsbad, CA) supplemented with 10% low antibody, heat-inactivated, gamma-irradiated fetal bovine serum (FBS, Hyclone, GE Healthcare Life Sciences, Pittsburgh, PA), L-alanine, L-glutamine dipeptide supplement (GlutaMAX,), and 50 μg/mL penicillin, 50 μg/mL streptomycin, 100 μg/mL neomycin (PSN antibiotics, Invitrogen) with incubation at 37°C in 5% CO_2_.

Attempts to isolate SARS-CoV-2 were performed in a BSL3 laboratory by an analyst who wore powered air-purifying respirators and used BSL3 work practices. Cells grown as monolayers in a T-25 flask (growing surface 25 cm^2^) were inoculated when they were at approximately 80% of confluency. First, aliquots (300 µL) of material extruded into VTM were filtered through a sterile 0.45 µm pore-size PVDV syringe-tip filter to remove bacterial and fungal cells and spores. Next, spent cell culture medium was removed and replaced with 1 mL of cell culture medium, and the cells inoculated with 50 μL of specimen filtrates. Post-inoculation, the cell cultures were incubated at 37°C in 5% CO_2_, and rocked every 15 minutes for 1 hour, after which 4 mL of complete cell growth medium with 3% FBS was added. Mock-infected cell cultures were maintained in parallel with the other cultures. The cell cultures were refed every three days by the replacement of 2 mL of spent media with complete aDMEM with 3% FBS. The cells were observed daily for one month before being judged negative for virus isolation. When virus-induced cytopathic effects (CPE) were evident, the presence of SARS-CoV-2 was determined by rRT-PCR [10].

SARS-CoV-2 genomic RNA (vRNA) was detected in culture media by real time reverse transcriptase polymerase chain reaction (rRT-PCR) following the procedure outlined in ref. 10. Briefly, vRNA was purified from spent cell culture media using a QIAamp Viral RNA Mini Kit (Qiagen, Valencia, CA, USA), with purified RNA eluted from the RNA-binding silicone column in a volume of 80 µL. rRT-PCR reactions were performed in a BioRad CFX96 Touch Real-Time PCR Detection System using 5 µL of purified vRNA and rRT-PCR primers that detect a section of the SARS-CoV-2 N-gene [11].

SARS-CoV-2 was isolated in all the cell lines tested, though compared to previous SARS-CoV-2 lineages we have isolated, virus-specific cytopathic effects (CPE) were much less pronounced and delayed by several days, in keeping with observations from other investigators [6-8]. In general, CPE were easiest to discern 6 or more days post inoculation. When examined by rRT-qPCR [10], the isolation of the SARS-CoV-2 omicron strains of this work was most effective in VeroE6-ACE-2 and HEKT293-ACE-2 cells (10^4^ to 10^7^ genome equivalents/µL of purified vRNA), with marginally lower virus yields in LLC-MK2 and VeroE6 cells, and generally low yields in A549-ACE-2 cells (10^1^ to 10^2^ genome equivalents/µL of purified vRNA).

***Statistical Analysis:*** The exact transition time from positive to negative was not observable for any child. We assume the transition occurred between the last positive test and the first subsequent negative test. This corresponds to interval censoring in the survival setting. For some children, there was no subsequent negative test after the last positive one, which corresponds to the right censoring. All children had at least one positive test which could be the initial diagnostic test performed before enrollment (i.e., outside this study). We fitted a parametric survival model to the data to account for interval and right censoring, with the time from symptom onset to the positive-to-negative transition as the time-to-event outcome. We made two assumptions: (1) every case would be positive if tested on the symptom onset day; and (2) the time from symptom onset to transition follows a Weibull distribution. There were three children with symptom onset dates missing. As the lag from symptom onset to the initial diagnostic test varied from 0 to 2 days with a median of 1 day for the majority of children (19 of 28) who had symptom onset dates available, we assume the symptom onset dates were one day before their initial diagnostic tests for the three children. We report the mean and median times from onset to transition and plotted the survival curve to show the changes in the percent of positive over time. Confidence intervals were calculated using the delta method. The survival model was fitted using the survival package in the statistical software R version 4.1.3 (R Core Team 2022).

**REFERENCES (Supplementary Methods)**

1. Shankar SN, Witanachchi CT, Morea AF, et al. SARS-CoV-2 in resdiential rooms of two self-isolating persons with COVID-19. J Aerosol Science 2022;159:1058701. <https://doi.org/10.1016/j.jaerosci.2021.105870>.

2. CDC. National Center for Immunization and Respiratory Diseases (NCIRD), Division of Viral Diseases - https://www.cdc.gov/coronavirus/2019-ncov/lab/rt-pcr-panel-primer-probes.html/. June 6, 2020.

3. Magalis BR, Rich S, Tagliamonte MS, *et al.* SARS-CoV-2 Delta vaccine breakthrough transmissibility in Alachua County, Florida. *Clin Infect Dis* 2022 Mar 10;ciac197. doi: 10.1093/cid/ciac197

4. Moshiri N. ViralMSA: Massively scalable reference-guided multiple sequence alignment of viral genomes. *Bioinformatics* 2020. doi:10.1093/bioinformatics/btaa743.

5. Rambaut A, Holmes EC, O’Toole A, *et al.* A dynamic nomenclature proposal for SARS-CoV-2 lineages to assist genomic epidemiology. *Nat Microbiol* 2020;5:1403-1407, doi:10.1038/s41564-020-0770-5 (2020).

6. Nguyen LT, Schmidt HA, von Haeseler A, Minh BQ. IQ-TREE: a fast and effective stochastic algorithm for estimating maximum-likelihood phylogenies. *Mol Biol Evol* 2015;32:268-274, doi:10.1093/molbev/msu300.

7. Zhao H, Lu L, Peng Z, *et al*. SARS-CoV-2 Omicron variant shows less efficient replication and fusion activity when compared with Delta variant in TMPRSS2-expressed cells, Emerging *Microbes & Infections* 2022;11(1):277-283, DOI: 10.1080/22221751.2021.2023329

8. Yadav PD, Gupta N, Potdar V, *et al*. Isolation and Genomic Characterization of SARS-CoV-2 Omicron Variant Obtained from Human Clinical Specimens. *Viruses* 2022;14(3):461. <https://doi.org/10.3390/v14030461>

9. Suzuki R, Yamasoba D, Kimura I *et al*. Attenuated fusogenicity and pathogenicity of SARS-CoV-2 Omicron variant. *Nature* (2022). <https://doi.org/10.1038/s41586-022-04462-1>

10. Lednicky JA, Lauzard M, Fan ZH, *et al*. Viable SARS-CoV-2 in the air of a hospital room with COVID-19 patients. *Int J Infect Dis.* 2020 Nov;100:476-482. doi: 10.1016/j.ijid.2020.09.025. Epub 2020 Sep 16. PMID: 32949774; PMCID: PMC7493737.

11. Lednicky JA, Shankar SN, Elbadry MA, *et al*. Collection of SARS-CoV-2 virus from the air of a clinic within a university student health care center and analyses of the viral genome. Aerosol Air Qual Res 20 (2020), pp. 1167–1171.

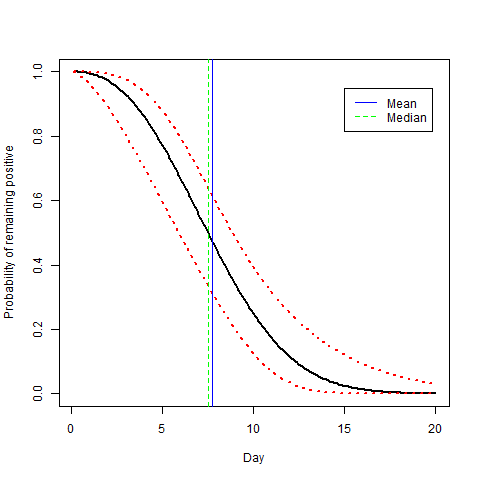

**Figure S1.** Model-predicted probability of remaining test-positive since symptom onset. This probability curve (black solid) is simply the survival function for the fitted Weibull model, and the 95% confidence bands (red dashed) are derived using the delta method. Mean (blue solid) and median (green dashed) times from symptom onset to turning negative are shown as vertical lines.
